# Strain measures of the left ventricle and left atrium are composite measures of left heart size and function

**DOI:** 10.1101/2023.05.04.23289077

**Authors:** Fredrika Fröjdh, Dhnanjay Soundappan, Peder Sörensson, Andreas Sigfridsson, Eva Maret, Jannike Nickander, Martin Ugander

## Abstract

**Background:** Left ventricular (LV) global longitudinal strain (GLS) and circumferential strain (GCS), and left atrial (LA) strain (LAS) are indicators of poor prognosis. However, it is unclear how they relate to each other and to LV and LA geometry. The aim of our study was to clarify the relationship between strain and LV and LA geometry to inform clinical and research applications.

**Methods:** Patients referred for cardiovascular magnetic resonance imaging were retrospectively identified. Univariable and multivariable linear regression models evaluated associations between GLS, GCS, LAS, LV mass, the volumes and dimensions of the LV and LA, and mitral annular plane systolic excursion (MAPSE).

**Results:** In patients (n=66, median [interquartile range] age 62 [53–72] years, 82% male, LV ejection fraction 48 [34–56]%, range 6–69%), GLS associated with both GCS (R^2^=0.86, p<0.001) and LAS (R^2^=0.51, p<0.001), and LAS associated with GCS (R^2^=0.42, p<0.001). GLS, GCS, and LAS were all univariably associated with MAPSE, LV mass, and the volumes and dimensions of the LV and LA (p<0.001 for all). In multivariable analysis, GLS associated with MAPSE and LV length (R^2^=0.85, p<0.001); GCS with MAPSE, LV end-systolic volume, and LV mass (R^2^=0.80, p<0.001); and LAS with LA end-diastolic volume and MAPSE (R^2^=0.67, p<0.001).

**Conclusions:** LV and LA strains are geometrically coupled composite measures of MAPSE and of LV and LA size and function, respectively. The composite of these geometric relationships likely explains the excellent prognostic association observed with strain measures. GLS and GCS provide largely overlapping information and should not be treated as distinct measures of longitudinal and circumferential function.

## Introduction

Deformation analysis (strain) from cardiac imaging of the left ventricle (LV), measured as global longitudinal strain (GLS) or global circumferential strain (GCS), and strain of the left atrium (LA), measured as left atrial strain (LAS), are increasingly used for assessment of LV function and patient prognosis. GLS and GCS are more sensitive^1^ and reproducible^2^ parameters of LV systolic function compared to LV ejection fraction (LVEF), providing incremental prognostic value beyond LVEF^3^. While LVEF is of importance in the characterization of heart failure (HF) according to clinical paradigms, LV strain is a more accurate measure for detecting the presence of LV dysfunction and evaluating disease progression over time^4,5^ as well as identifying compromised LV systolic function in patients with preserved EF^6^. Moreover, the analysis of left atrial function using left atrial strain, LAS, also referred to as LA reservoir strain, is emerging as a means for non-invasive assessment of diastolic dysfunction. LAS demonstrates good agreement with the degree of diastolic function as assessed by 2016 American Society of Echocardiography guidelines^7^ as well as with invasive pressure measurements. As opposed to many other parameters used to assess diastolic dysfunction, LAS correlates linearly with the severity of disease and has been suggested a potential supplementary or stand-alone parameter for the non-invasive assessment of LV filling pressure^8–10^. However, the physiologic basis for this association remains poorly understood. Alternating and reciprocal shortening and lengthening of the LV and LA, respectively, is intrinsically coupled through the shared atrioventricular valve (AV) plane. Notably, the LV apex and the posterior aspect of the LA remain effectively stationary throughout the cardiac cycle, and as a result, longitudinal movement of the LA and LV almost exclusively translates to the displacement of the AV plane, measured as mitral annular plane systolic excursion (MAPSE). Consequently, MAPSE correlates with GLS, and this correlation increases when MAPSE is indexed to LV length, providing similar prognostic strength compared with GLS^11,12^. By analogy, we hypothesize that LAS is mainly explained by the variation in MAPSE and left atrial size and that the correlation between LAS and GLS is driven by these associations.

Thus, the aim of this study was to determine the correlation between LA and LV strain, LA and LV dimensions, and MAPSE, with the purpose of better understanding these commonly used measures in both cardiovascular research and clinical cardiac imaging.

## Methods

### Population

We retrospectively screened patients aged 18 or older, who had undergone clinically indicated cardiovascular magnetic resonance imaging (CMR) between December 2017 and May 2021 and had provided written informed consent. Ethical approval was obtained from the Regional Ethics Committee in Stockholm, Sweden (Approval number Dnr 2011:1077-31:3), and the study was conducted in accordance with the principles outlined in the Declaration of Helsinki. The population was selected to represent a variable spectrum of LVEF, including 20 – 25 patients from each of the following ranges: LVEF <30%, 30 – 40%, 40 – 50%, 50 – 60%, and >60%, while excluding those with distinct myocardial pathologies such as hypertrophic cardiomyopathy, amyloidosis, hemosiderosis, Anderson-Fabry’s disease, congenital heart defects, atrial fibrillation during CMR, or insufficient image quality to perform LV strain analysis. Further exclusion criteria included inadequate tracking of the myocardium by strain image analysis, defined as visually inadequate tracking of >2 segments in any of the long-axis or short-axis views, and visually apparent foreshortening or the presence of ≥3 of the atrial appendage and pulmonary veins in both 2- and 4-chamber views.

### CMR imaging

Clinical CMR scans were performed at 1.5T or 3T MAGNETOM (Aera or Skyra, Siemens Healthcare, Erlangen, Germany) with an 18-channel and 32-channel phased-array body and spine coil, respectively. The exam included retrospectively ECG-gated balanced steady-state free precession cine imaging in short axis (8 mm slice thickness, 1.6 mm slice gap) and long axis 2-, 3-, and 4-chamber views (8 mm slice thickness) with 30 phases per cardiac cycle.

### Image analysis

All post-processing analyses were performed using freely available software Segment version 3.0 R9405e (Medviso AB, Lund, Sweden). LV volumes were assessed by manual delineation of the endo- and epicardium according to guidelines ^13^, excluding papillary muscles for LV mass assessment. LV GLS and LV GCS were separately analyzed using a semi-automated feature-tracking module using cine images by an observer (FF) blinded to clinical data and geometric measures. Endo- and epicardial borders were manually traced in end-diastole and automatically propagated throughout the cardiac cycle. LV GLS was measured as the mean longitudinal strain in two-, three- and four-chamber views, and LV GCS as the mean circumferential strain in an apical, midventricular, and basal short-axis view. Delineations were adjusted manually if ≥2 segments did not track myocardial motion adequately as assessed visually.

LA dimensions and volumes were measured using the bi-plane area-length method in the two- and four-chamber views by a single-blinded observer (DS), excluding the left atrial appendage and pulmonary veins. LAS was assessed as the strain measured from end diastole to end systole in a two- or four-chamber view or the mean of both. LV and LA measurements were performed by two different observers.

MAPSE was measured as the mean distance in millimeters traveled by the mitral annular insertion points from end-diastole to end-systole, averaged from two manual caliper measurements per view in the 2- and 4-chamber views. In the 3-chamber view, the lateral mitral annular insertion point and septal aortic valve insertion point were used. LV length (LVL) was measured as the distance from the most apical point of the epicardium to the midpoint between the mitral annular insertion points in end-diastole. The longitudinal contribution to LV stroke volume was calculated as the maximal LV short-axis epicardial area multiplied by MAPSE as previously described and validated ^14^.

### Statistical analysis

Statistical analysis was performed using RStudio Team (2022). Patient characteristics were summarized as the median [interquartile range] for continuous variables and as counts and percentages for categorical variables. Univariable linear regression was used to assess correlations. Multivariable linear regression was used to identify measures associated with GLS, GCS, and LAS, excluding the variable with the lowest univariable correlation in the case of high multicollinearity. Agreements between GCS and GLS were assessed using Bland-Altman plots. A p-value less than 0.05 was considered statistically significant.

## Results

### Patient characteristics

A flowchart of patient selection is shown in Figure 1. The final study population included 66 patients: 14 (18%) were female, age was 62 [53 – 72] years and LVEF was 48 [34 – 57]%, full range 9 – 69%. Characteristics of the study population are shown in Table 1.

**Figure 1.**
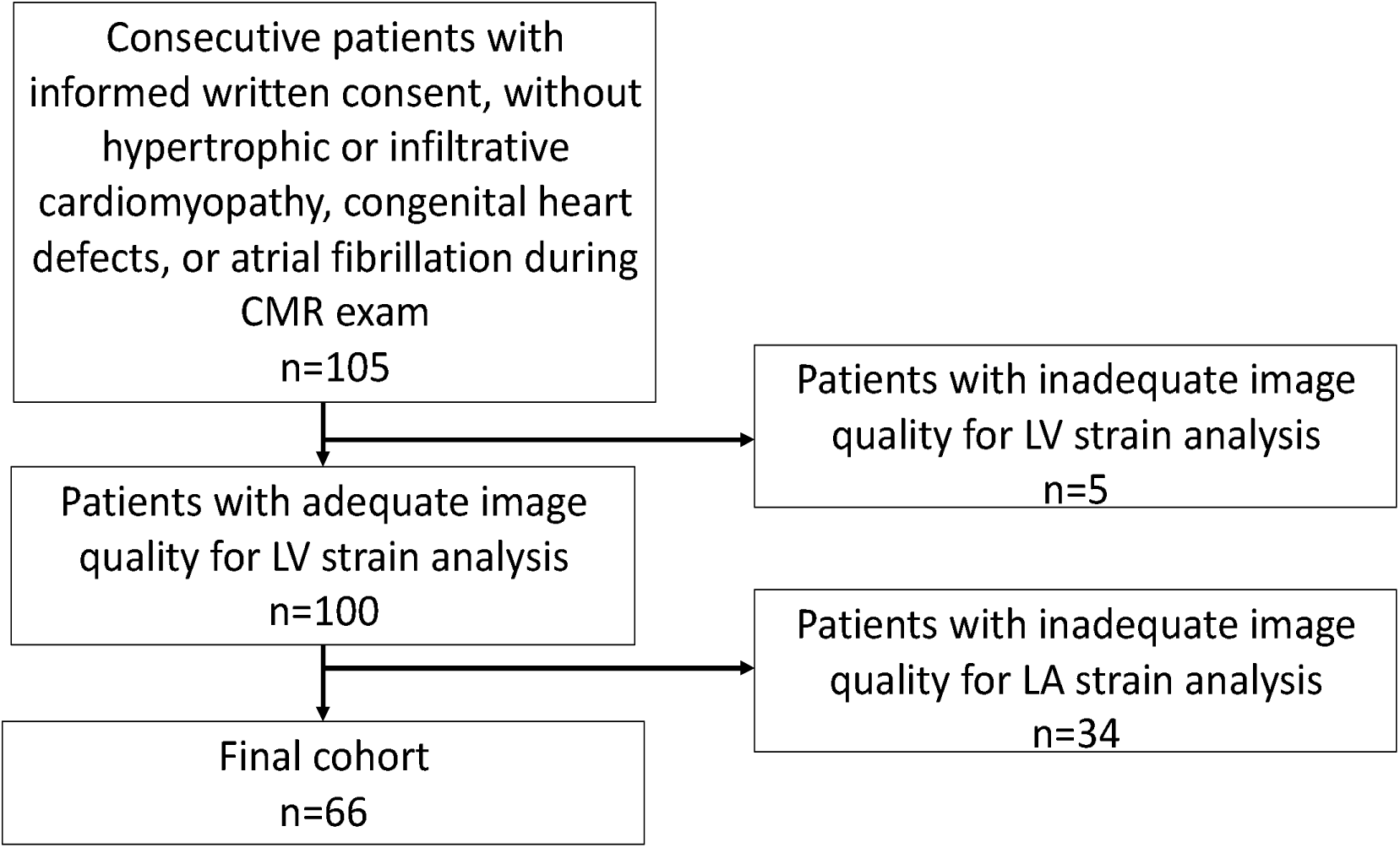
Flowchart of patient inclusion

**Table 1.**
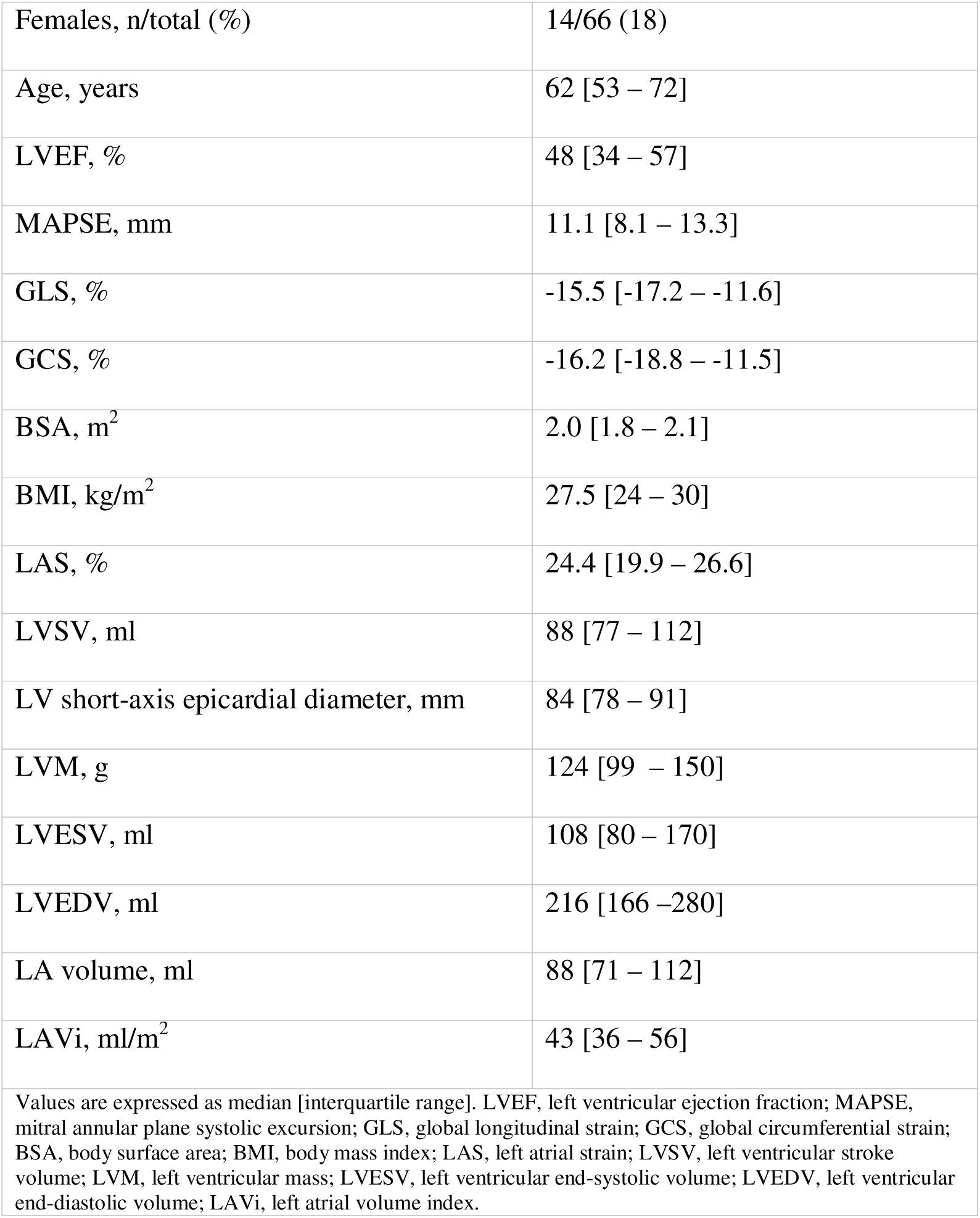
Baseline characteristics.

|  |  |
| --- | --- |
| Females, n/total (%) | 14/66 (18) |
| Age, years | 62 [53 – 72] |
| LVEF, % | 48 [34 – 57] |
| MAPSE, mm | 11.1 [8.1 – 13.3] |
| GLS, % | -15.5 [-17.2 – -11.6] |
| GCS, % | -16.2 [-18.8 – -11.5] |
| BSA, m<sup>2</sup> | 2.0 [1.8 – 2.1] |
| BMI, kg/m<sup>2</sup> | 27.5 [24 – 30] |
| LAS, % | 24.4 [19.9 – 26.6] |
| LVSF, ml | 88 [77 – 112] |
| LV short-axis epicardial diameter, mm | 84 [78 – 91] |
| LVM, g | 124 [99 – 150] |
| LVESV, ml | 108 [80 – 170] |
| LVEDV, ml | 216 [166 – 280] |
| LA volume, ml | 88 [71 – 112] |
| LAVi, ml/m<sup>2</sup> | 43 [36 – 56] |
| Values are expressed as median [interquartile range]. LVEF, left ventricular ejection fraction; MAPSE, mitral annular plane systolic excursion; GLS, global longitudinal strain; GCS, global circumferential strain; BSA, body surface area; BMI, body mass index; LAS, left atrial strain; LVSF, left ventricular stroke volume; LVM, left ventricular mass; LVESV, left ventricular end-systolic volume; LVEDV, left ventricular end-diastolic volume; LAVi, left atrial volume index. |  |

### Global longitudinal strain

A summary of the relationships between strain values and geometric measures is illustrated in representative patients in Figure 2. GLS was -15.5% [-17.2 – -11.6] in the study population. GLS was highly correlated with GCS (R^2^ = 0.86, p <0.001). GLS was also highly correlated with conventional LV metrics, including LV mass, EDV, ESV, LV diameter, MAPSE, and LV length, but not with BSA or age. In multivariable regression analysis using GLS as the dependent variable, MAPSE, LV mass, and LV ESV together contributed to the model with the strongest association (R^2^ = 0.89, p <0.001). MAPSE modeled together with other parameters of LV size yielded models of similar strength of association (MAPSE and LVEDV, R^2^ = 0.87, p <0.001; MAPSE and LV length, R^2^ = 0.84, p <0.001; MAPSE and LV maximal diameter, R^2^ = 0.85, p <0.001). In these multivariable models, the association with LV mass did not remain significant. Moreover, LVEDV and LVM were correlated (R^2^ = 0.51, p <0.001). Results from univariable and multivariable linear regression analysis are presented in Table 2. GLS was strongly associated with MAPSE indexed to LV length (R^2^ = 0.84 p <0.001) and with MAPSE indexed to the sum of LV length and LV diameter (R^2^ = 0.84, p <0.001). GLS could be estimated with high accuracy and precision from MAPSE indexed to LV length (difference 3.7 ± 1.9 %-points). The linear relationship between MAPSE/LVL and GLS was expressed as (Figure 3):

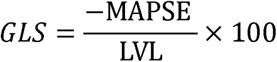

**Figure 2.**
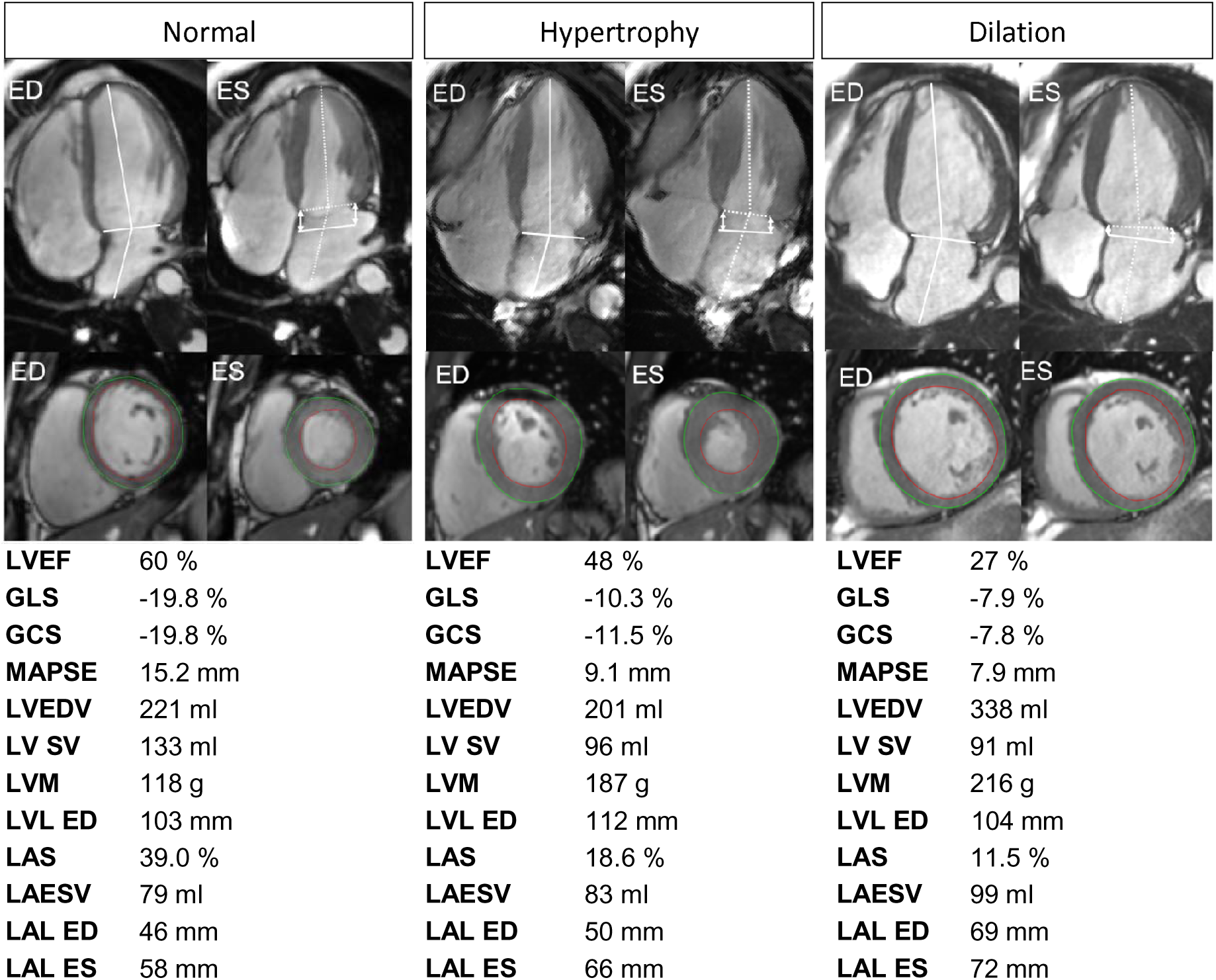
Representative cases highlighting the association between strain and geometric measures. All images are shown in the same magnification. End-diastolic (ED) and end-systolic (ES) long-axis (top row) and short-axis (lower row) images of three representative patients with normal (left), hypertrophied (middle), and dilated (right) geometry shown together with measures of left ventricular ejection fraction (LVEF), global longitudinal strain (GLS), global circumferential strain (GCS), mitral annular plane systolic excursion (MAPSE), LV mass (LVM) and end-diastolic volume (EDV), stroke volume (SV), left atrial strain (LAS), left atrial end-systolic volume (LAESV) and left atrial length (LAL). The white lines denote the LV and left atrial length (LAL) in ED and the white dotted lines LV and LA length in ES. The white double-headed arrows denote septal and lateral MAPSE, respectively. Across all patients and magnitudes of LV function, approximately 60% of the LV stroke volume is generated by the longitudinal movement of the atrioventricular plane (MAPSE), that simultaneously empties and fills the LV and LA. Thus, MAPSE mechanically couples the function of the LV and LA to each other. Meanwhile, the LV apex and the posterior aspect of the LA are effectively stationary throughout the cardiac cycle. Note that across these representative patients, 1) Intra-individual GLS and GCS are very similar, 2) GLS is a function of both MAPSE and LV volume or length, and 3) LAS is a function of MAPSE and LA volume or length.

**Figure 3:**
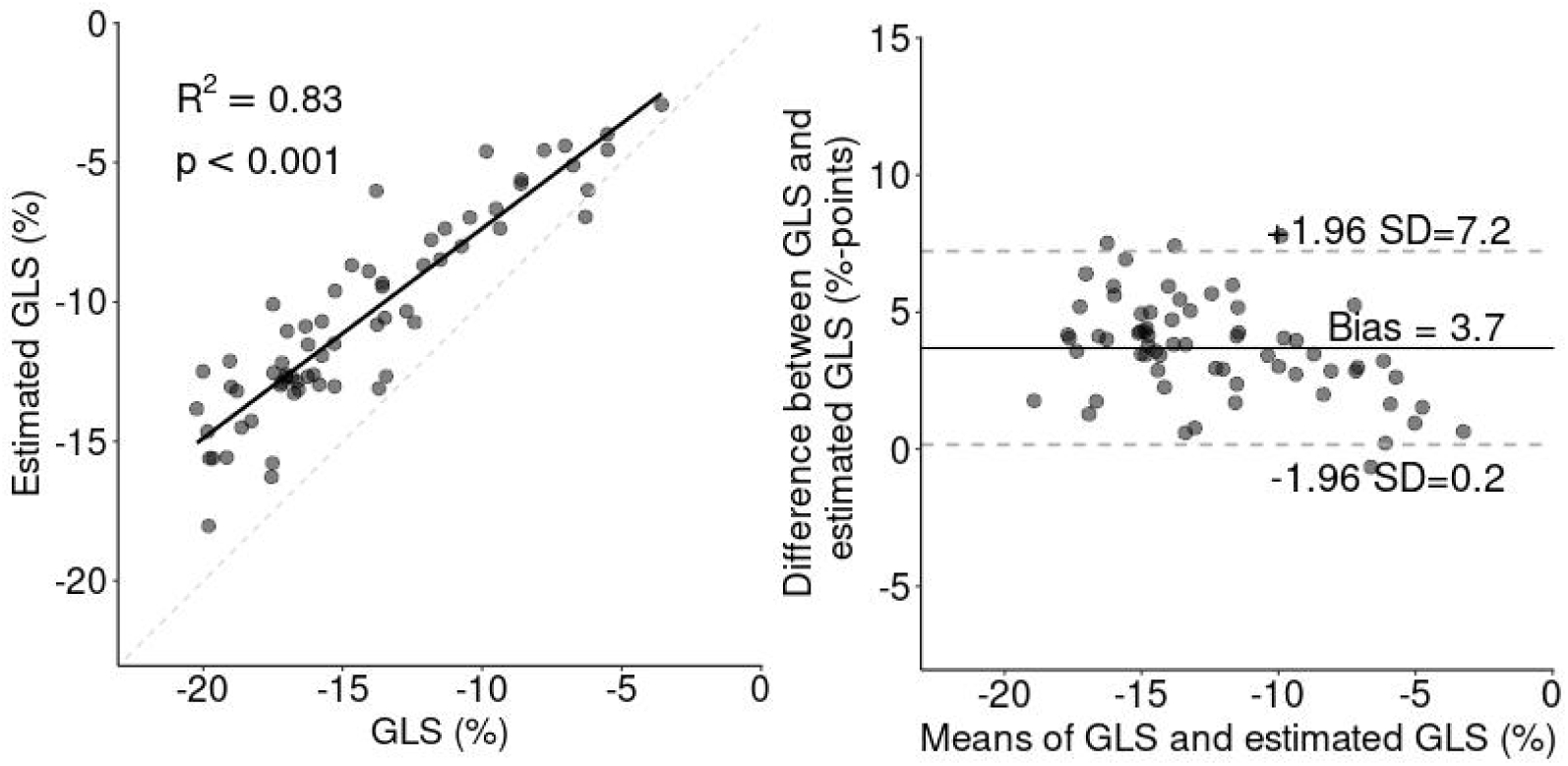
Agreement between measured GLS and estimated GLS using MAPSE and LV length. Left: Linear regression (solid line) plot showing the linear correlation between estimated global longitudinal strain (GLS) and GLS, y= -1.10x + 2.59. The dashed line indicates the line of identity. Right: Bland-Altman plot showing the agreement between GLS and GLS estimated as *-*MAPSE/LVL *\*100*. Bias±SD was 3.69 ± 1.87%.

**Table 2.** Associations between GLS and left heart measures.

|  | Univariable |  | Multivariable |  |  |
| --- | --- | --- | --- | --- | --- |
| | R<sup>2</sup> | p | Standardized $\beta$ | p | Global R<sup>2</sup> |
| LVEF, % | 0.82 | **<0.001** |  |  | 0.89<br>**p<0.001** |
| GCS, % | 0.86 | **<0.001** |  |  |  |
| MAPSE, mm | 0.78 | **<0.001** | -0.59 | **<0.001** |  |
| Indexed MAPSE, %, | 0.83 | **<0.001** |  |  |  |
| LVL, mm | 0.23 | **<0.001** |  |  |  |
| SV, ml | 0.19 | **<0.001** |  |  |  |
| SA area, cm<sup>2</sup> | 0.53 | **<0.001** |  |  |  |
| SA area cm<sup>2</sup> × MAPSE/SV | 0.01 | 0.69 |  |  |  |
| LVM, g | 0.33 | **<0.001** | 0.12 | **0.04** |  |
| LVEDV, ml | 0.50 | **<0.001** |  |  |  |
| LVESV, ml | 0.69 | **<0.001** | 0.37 | **<0.001** |  |
| LAS, % | 0.51 | **<0.001** |  |  |  |
| LA, ml | 0.13 | **<0.01** |  |  |  |

### Global circumferential strain

GCS was -16.2% [-18.75 – -11.6] in the study population. Similarly to GLS, GCS was strongly correlated with LV mass, stroke volume, LVEDV, LVESV, MAPSE, LV length, LVEF, and LV diameter, but not with BSA (Table 3). In multivariable analyses, LVESV and MAPSE together produced the best model for estimating GCS.

**Table 3.**
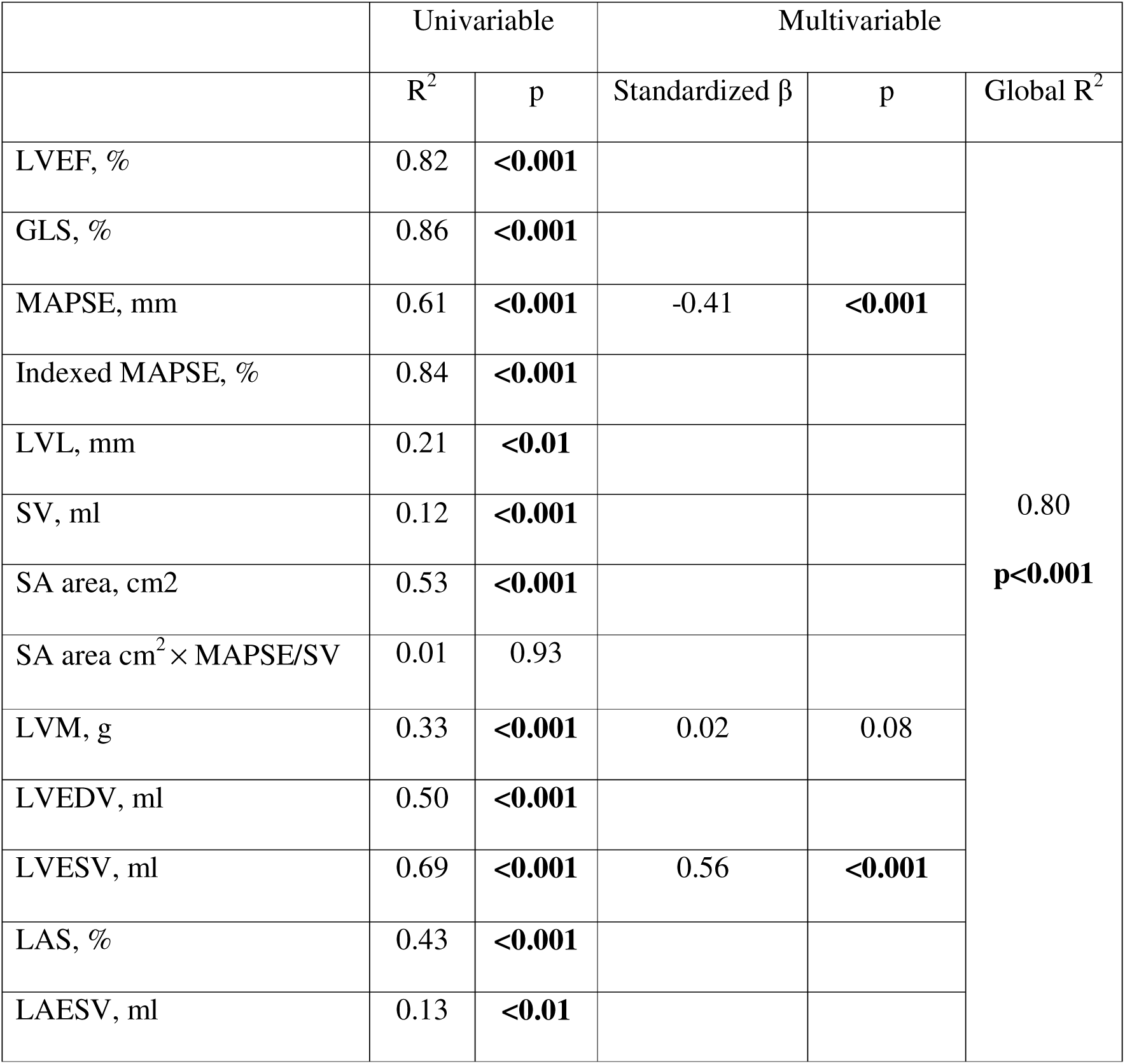
Associations between GCS and left heart measures.

|  | Univariable |  | Multivariable |  |  |
| --- | --- | --- | --- | --- | --- |
| | R<sup>2</sup> | p | Standardized $\beta$ | p | Global R<sup>2</sup> |
| LVEF, % | 0.82 | **<0.001** |  |  | 0.80<br>**p<0.001** |
| GLS, % | 0.86 | **<0.001** |  |  |  |
| MAPSE, mm | 0.61 | **<0.001** | -0.41 | **<0.001** |  |
| Indexed MAPSE, % | 0.84 | **<0.001** |  |  |  |
| LVL, mm | 0.21 | **<0.01** |  |  |  |
| SV, ml | 0.12 | **<0.001** |  |  |  |
| SA area, cm<sup>2</sup> | 0.53 | **<0.001** |  |  |  |
| SA area cm<sup>2</sup> × MAPSE/SV | 0.01 | 0.93 |  |  |  |
| LVM, g | 0.33 | **<0.001** | 0.02 | 0.08 |  |
| LVEDV, ml | 0.50 | **<0.001** |  |  |  |
| LVESV, ml | 0.69 | **<0.001** | 0.56 | **<0.001** |  |
| LAS, % | 0.43 | **<0.001** |  |  |  |
| LAESV, ml | 0.13 | **<0.01** |  |  |  |

### Left atrial strain

LAS, measured as LA reservoir strain, was 24 [20 – 27] % in the study population. Results from the linear analyses of LA and LV parameters are summarized in Table 4. LAS correlated with GLS (R^2^ = 0.51, p <0.001). MAPSE associated positively with LAS (R^2^ = 0.46, p <0.001), and this correlation increased when MAPSE was indexed to LA length (R^2^ = 0.51, p <0.001), in analogy to MAPSE indexed to LVL. In multivariable analysis, LAEDV and MAPSE together yielded the strongest association with LAS (R^2^ = 0.67, p <0.001).

**Table 4.**
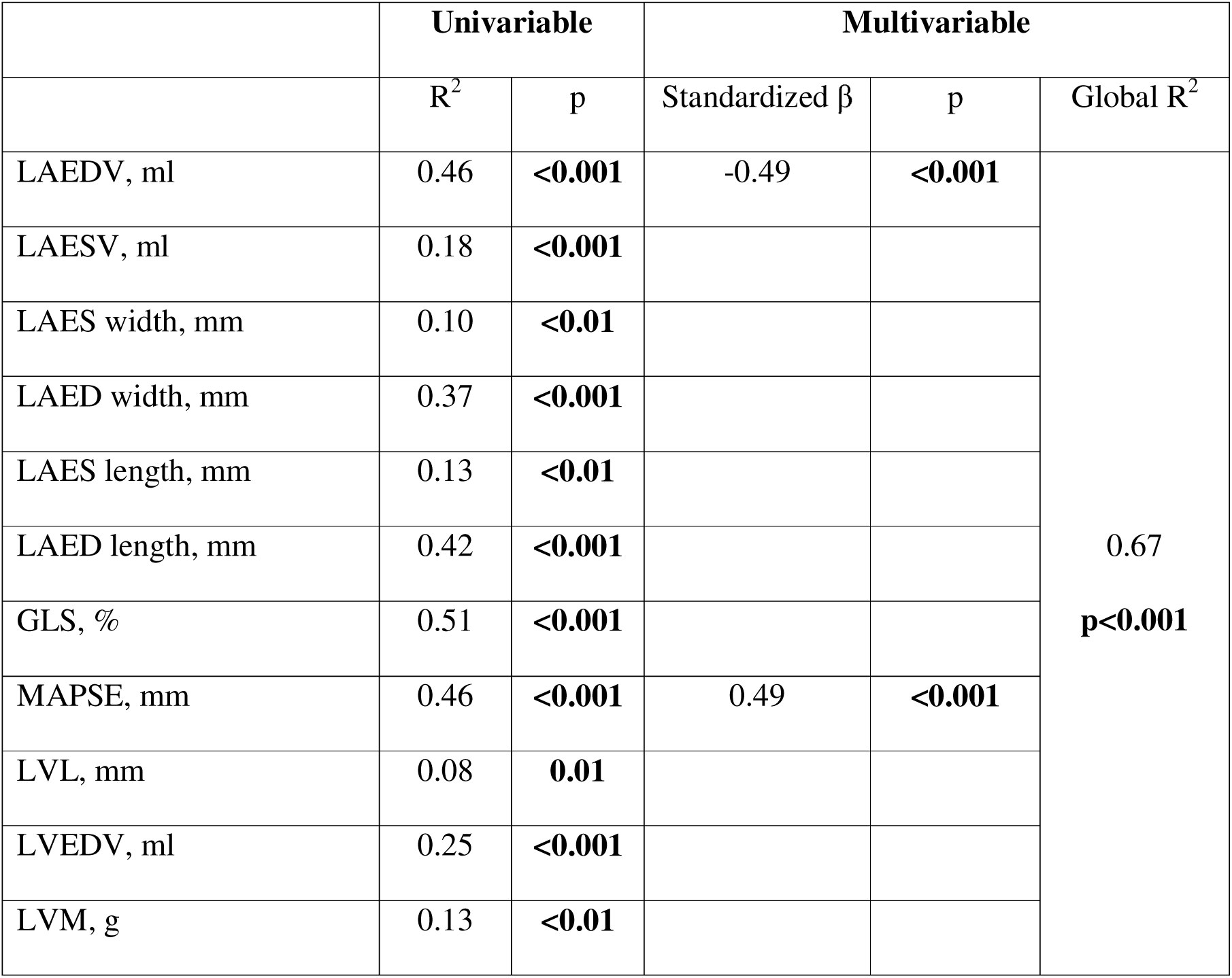
Associations between LAS and left heart measures.

|  | Univariable |  | Multivariable |  |  |
| --- | --- | --- | --- | --- | --- |
| | R<sup>2</sup> | p | Standardized $\beta$ | p | Global R<sup>2</sup> |
| LAEDV, ml | 0.46 | **<0.001** | -0.49 | **<0.001** | 0.67<br>**p<0.001** |
| LAESV, ml | 0.18 | **<0.001** |  |  |  |
| LAES width, mm | 0.10 | **<0.01** |  |  |  |
| LAED width, mm | 0.37 | **<0.001** |  |  |  |
| LAES length, mm | 0.13 | **<0.01** |  |  |  |
| LAED length, mm | 0.42 | **<0.001** |  |  |  |
| GLS, % | 0.51 | **<0.001** |  |  |  |
| MAPSE, mm | 0.46 | **<0.001** | 0.49 | **<0.001** |  |
| LVL, mm | 0.08 | **0.01** |  |  |  |
| LVEDV, ml | 0.25 | **<0.001** |  |  |  |
| LVM, g | 0.13 | **<0.01** |  |  |  |

### Longitudinal and circumferential contribution to left ventricular stroke volume

The longitudinal contribution to LV stroke volume was 62 [58 – 66]% in the population. The longitudinal contribution to LV stroke volume did not associate with GLS (p = 0.69) or MAPSE indexed to LV length (p = 0.12). Similarly, GCS was not associated with the short-axis contribution to LV stroke volume (p = 0.93).

### Relationship between MAPSE and LV and LA geometries

MAPSE was univariably correlated with LV and LA strain as detailed above, as well as with LV EDV, (p <0.001), LV ESV (p = 0.01), LVM (p <0.001) LVEF (p <0.001), LA EDV (p < 0.001), and trended to a correlation with LA ESV (p = 0.06), see Table 5. In multivariable analysis excluding strain measures for multicolinearity reasons, only LVESV remained associated.

**Table 5.** Associations between MAPSE and left heart measures.

|  | Univariable |  | Multivariable |  |  |
| --- | --- | --- | --- | --- | --- |
| | R<sup>2</sup> | p | Standardized $\beta$ | p | Global R<sup>2</sup> |
| LAEDV, ml | 0.22 | **<0.001** |  |  | 0.61<br>**p<0.001** |
| LAESV, ml | 0.05 | **0.06** | 0.13 | 0.20 |  |
| GLS, % | 0.78 | **<0.001** |  |  |  |
| EF, % | 0.71 | **<0.001** |  |  |  |
| LAS, % | 0.57 | **<0.001** |  |  |  |
| LVM, g | 0.17 | **<0.001** | -0.44 | **0.003** |  |
| LVL, mm | 0.06 | **0.03** | 0.72 | **<0.001** |  |
| SA area cm<sup>2</sup> | 0.33 | **<0.001** |  |  |  |
| LVESV, ml | 0.45 | **0.01** | -0.97 | **<0.001** |  |
| LVEDV, ml | 0.25 | **<0.001** |  |  |  |
| SV, ml | 0.37 | **<0.001** |  |  |  |
| Age | 0.01 | **0.64** |  |  |  |

## Discussion

We found that LV strain strongly associated with MAPSE and LV dimensions, representing a composite measure of both function and LV geometry. Similarly, LA strain was closely associated with MAPSE and LA volume. All strain measures were highly correlated, offering insights that aid clinical and clinical research interpretation by considering the associations between strain measures and conventional geometric measures of LV and LA size and function.

### GLS

GLS was closely correlated with MAPSE, with the strength of this association increasing when MAPSE was indexed to LV length (R^2^ = 0.83), of similar degree compared to previous published CMR (R^2^ = 0.85)^11^ and echocardiographic data (R^2^ = 0.79)^15^. This confirms the dependence of GLS on atrioventricular plane displacement and LV size. Additionally, GLS could be accurately and precisely estimated from MAPSE and LV length using a simple linear formula, with similar accuracy compared to that found in a study using transthoracic echocardiographic data in 220 patients (bias 3.0 ± 3.5 vs 0.0 ± 4.8 %-points)^15^. The difference in bias between the methods can be attributed to the distinct measurement modalities used. Specifically, MAPSE with M-mode is measured at an angle with respect to the LV longitudinal axis, incorporating both longitudinal and some radial movement, whereas MAPSE with CMR is measured along the longitudinal axis alone. The slightly lower variation in the CMR data is likely due to GLS and MAPSE being measured within the same image. Conversely, echocardiographic MAPSE and speckle tracking data are typically acquired at different times during the examination. Given the sizeable inter-vendor variability in strain measures^16^, estimating GLS using MAPSE indexed to LV length offers a simple, vendor-independent, and reliable method. In contrast to GLS, both MAPSE and LV length measurements are that are less sensitive to image quality and may serve as surrogate markers for GLS in cases of limited image quality. This is in line with the previous findings of two independent studies in which MAPSE indexed to LV length provided comparable prognostic value to GLS^11,12^.

GLS was also highly correlated with LVEF. Like LVEF, GLS measures the relative change in LV size, although LVEF represents the inner volume change, GLS represents the change in mid-wall circumference of the LV myocardium in the long-axis direction, and as a consequence, incorporates both LV volume and myocardial wall thickness. This means that in most patients, GLS and LVEF are concordant. However, in patients with increased myocardial wall thickness, the greater difference in endocardial and mid-wall circumference translates to a relatively lower GLS compared to LVEF. This depends on the degree of hypertrophy, and consequently GLS provides incremental functional and prognostic information beyond LVEF in such patients^17^.

GLS correlated with myocardial mass in our cohort. However, when considering other dimensional parameters, this correlation did not retain statistical significance, most likely due to multicollinearity with LV volume. This explanation may be particularly prominent in our cohort that excluded patients with hypertrophic cardiomyopathy.

*GCS*. GLS and GCS were highly correlated with each other. Although a direct correlation between circumferential and longitudinal strain has not previously been reported, both have been found to correlate linearly with LVEF, indirectly supporting their high degree of correlation^18^.

Strain measures are two-dimensional, simultaneously assessing both shortening along one direction and the resultant perpendicular myocardial thickening. It should be noted that myocardial mass does not change during the cardiac cycle, it rearranges. Radial inward motion is incorporated in both GLS and GCS measures. The high correlation between GLS and GCS can thus both be explained by their inherent anatomical interdependence and the fact that myocyte function most likely varies little depending on its location in the myocardium. GCS also showed a strong correlation with MAPSE, and this is not surprising given the multi-collinearity of the already described correlations. Similarly to GLS, the correlation with GCS increased when MAPSE was indexed to LV length or LV diameter. In summary, GLS and GCS are both measures of global LV function and should not be viewed as distinctly separate measures of longitudinal and circumferential function, respectively.

### Associations with longitudinal function

In the current study, the median longitudinal contribution to LV stroke volume was 62%, consistent with previous research showing that longitudinal function varies very little across a broad range of LVEF values^19^. The longitudinal contribution to stroke volume was not correlated with GLS or MAPSE and, similarly, GCS was not correlated with the short-axis contribution to LV stroke volume. Therefore, while MAPSE and GLS measure longitudinal shortening, they may not be appropriate to describe in terms of longitudinal function or the longitudinal contribution to stroke volume *per se*.

### LAS

Similar to LV strain, LAS was highly positively correlated with LV systolic longitudinal shortening (MAPSE) and inversely correlated with LA dimensions. The Graphical Abstract (figure 4) provides a schematic overview of the physiologic relationships between strain measures, MAPSE and geometry. Although the correlation between MAPSE, LA measures, and LA strain was not as strong as that seen for LV strain, this may be due to the more irregular shape of the LA and the insertion of pulmonary veins and the atrial appendage^24^. Subsequently, LAS was highly correlated with both GLS and GCS, corroborating previous findings^20^. Our results indicate that LAS to a substantial degree can be explained by the variation in MAPSE and LA dimensions. As systolic function is a common cause of increased filling pressures and LA dilatation is a well-recognized consequence of prolonged pressure overload^21^, these correlations provide a physiologic mechanism coupling LAS with increased filling pressures. Because of this, LAS can also be seen to function as a composite marker of reduced myocardial function and left atrial enlargement. While left atrial strain (LAS) may be influenced by the duration of elevated filling pressures, it is also influence by additional factors contributing to left atrial (LA) dilatation such as mitral valve dysfunction and intermittent atrial fibrillation. Consequently, the applicability of LAS in certain patient populations may be less reliable and warrants further study.

**Figure 4.**
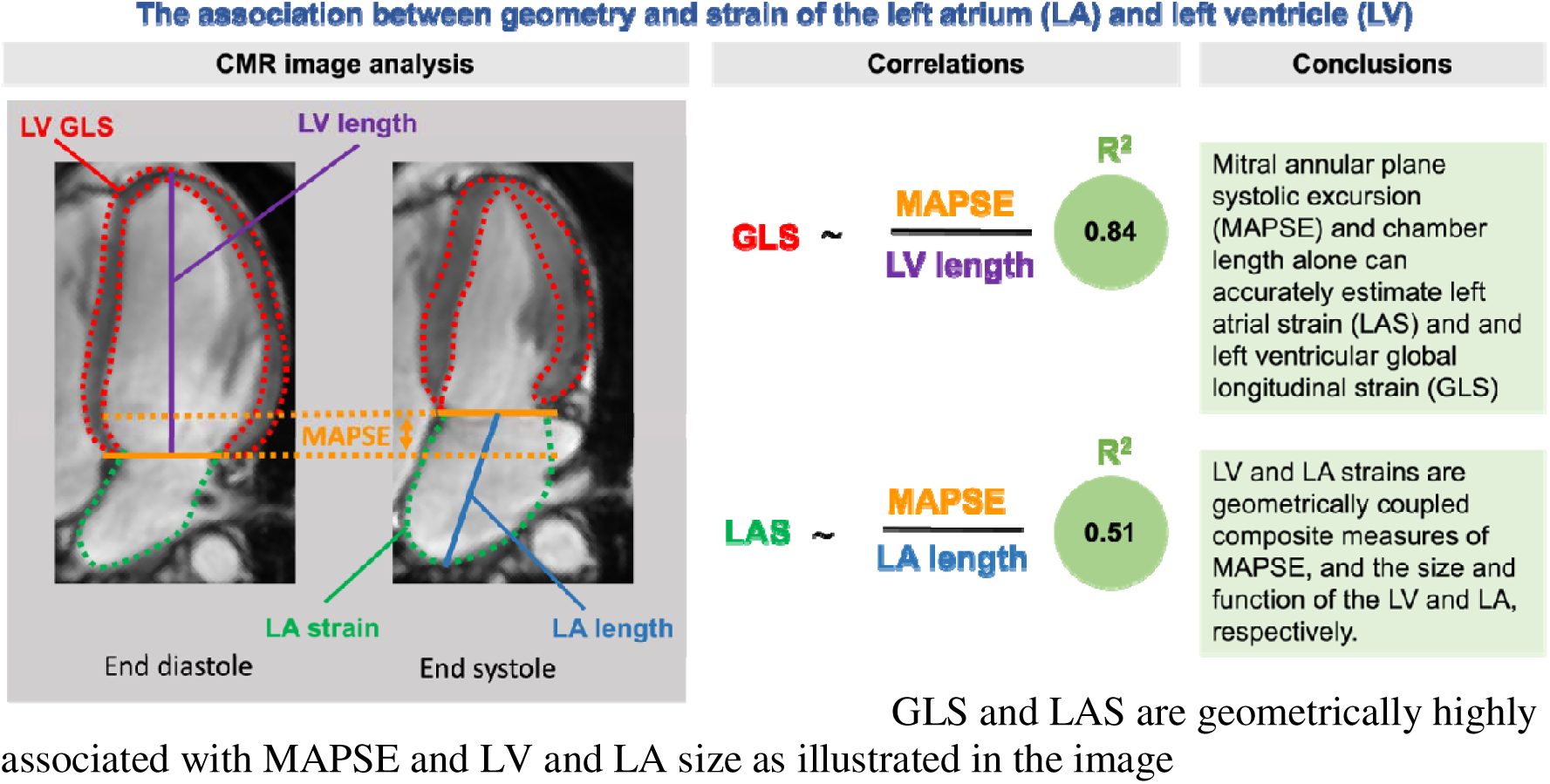
Graphical Abstract GLS and LAS are geometrically highly associated with MAPSE and LV and LA size as illustrated in the image

### MAPSE

MAPSE unsurprisingly correlated inversely with both LV volumes and myocardial mass, parameters associated with myocardial remodelling and loss of myocyte function, particularly in our cohort in the studied population mainly represented by patients with myocarditis or ischemic heart disease. Naturally, the relationships between MAPSE and LV dimensional measures would be expected to vary between different cardiac pathologies.

### Limitations

Images used for strain analyses were standard LV projections, and consequently optimized for visualization of the LV but not the LA. Given the irregular shape of the LA, pulmonary vein and atrial appendage insertion as well as a varying LA-to-LV angle, this likely affected LA length and volume calculations as well as LAS analyses and may in part account for the lower correlation between LAS and GLS-derived estimates of LAS compared to those found previously^22^. The majority of patients included in the current study had some degree of focal myocardial scarring as a consequence of including patients referred for CMR while excluding several other common clinical indications for CMR assessment including hypertrophic cardiomyopathy, valvular, and congenital heart disease. Focal scarring might bias strain measurements differently depending on the location in the myocardium. However, the exceptionally high correlation between GLS and GCS indicates that this potential source of variability is unlikely to have a large magnitude of effect on the findings. Also, infarct size associates with the magnitude of decrease in MAPSE, but not infarct location^23^. Thus, focal myocardial scarring should not represent a major limitation of this study.

The relative underrepresentation of females in our study may be seen as a limitation. However, while females are known to have smaller hearts than males on a group level, there is no indication that female and male hearts of the same size exhibit different proportions or geometric scaling. Hence, the results should be reasonably applicable across both sexes.

## Conclusions

LV and LA strain can be understood as geometrically coupled composite measures of MAPSE, and the size, function, and dimensions of the LV and LA. MAPSE and LV length alone can be used to accurately estimate GLS, which in turn provides similar information to GCS. These highly correlated composite geometrical relationships between strain and geometric measures of known prognostic significance likely explain the excellent prognostic value of strain measures. The research and clinical evaluation of various cardiac pathologies necessitates careful consideration of the dependence of strain measures on cardiac dimensions and myocardial function.

## Data Availability

All data produced in the present study are available upon reasonable request to the authors

## Authors contribution

F.F.: Conceptualization, methodology, investigation, formal analysis, writing – original draft.

M.U.: Conceptualization, methodology, supervision, data curation.

J.N.: Conceptualization, supervision, funding acquisition.

A.S.: Data curation.

D.S.: Methodology, data curation.

E.M.: Methodology, supervision.

P.S.: Methodology, supervision.

All authors contributed to writing – review and editing and approved the final version of the manuscript.

## Data availability

No additional data can be made available. Per the ethical approval, data can only be accessed locally via secure servers. This is to safeguard participants’ anonymity, and for this reasondata cannot be made generally available.

